# The role of digital systems in improving and strengthening routine Death Reporting in Civil Registration systems

**DOI:** 10.1101/2025.09.09.25335397

**Authors:** Collins Ochieng Odhiambo, Esther Nyadzua Katama, Brian Munkombwe, Chomba Mwango, Saviour Moyo, Chabila Mapoma

## Abstract

**Background:** Enhancing both notification and registration practices is crucial for generating stronger evidence to guide population health policy and planning. Strengthening civil registration systems for death reporting is therefore a vital first step toward improving the evidence base for effective health interventions.

**Methods:** This case study for Zambia examines how integrating digital and mobile technologies into death reporting can improve timeliness, efficiency, and accuracy compared to traditional manual practices. The study focused on burial offices in high-mortality provinces and districts, where improvements could yield substantial gains in reporting completeness. An electronic version of Form 13, the official death notification form, was developed to capture facility and community-based deaths in real time, before mortuary admission. A dashboard was created to track data collection progress, provide real-time access, and support analysis for policy action.

**Results:** Over the course of six months, the electronic platform captured approximately 100,000 deaths, with daily data streaming. Children under one year accounted for the largest proportion of deaths reported (18,884; 16.34%), while deaths among those aged 5–9 (1,977; 1.7%) and 10–14 (1,758; 1.5%) were the least common. The electronic system reduced reporting delays by 80% (e-reporting: 3 days; manual system: 24 days; p < 0.001). Completing a form electronically took an average of 4.6 minutes (median: 4.0).

**Conclusions:** Digitizing CRVS death reporting in Zambia significantly improved the timeliness, completeness, and quality of mortality data, enabling rapid analysis and policy response. The system also eased administrative burdens for bereaved families by reducing delays and unnecessary steps in the reporting process. These findings highlight the potential of digital platforms to strengthen death reporting, improve disease surveillance, and support data-driven interventions.

## Introduction

Civil registration and vital statistics (CRVS) death reporting and notification are essential for accurately documenting mortality in any country. Reliable CRVS records and the resulting statistics underpin national health and population policies, enabling governments to respond to emerging epidemics (1). Despite this importance, the World Health Statistics reveal that many low– and middle-income countries (LMICs) have low death registration rates and or produce incomplete or unreliable vital statistics (2).

Strengthening death registration is therefore a critical public health priority. The United Nations Sustainable Development Goal 16 (3, 4) explicitly targets the improvement of CRVS systems as part of enhancing human security.

In well-developed CRVS systems, digitization supports the collection of high-quality, timely mortality data and facilitates prompt notification of deaths (5, 6). However, many developing countries—such as Zambia—face a higher burden of disease and mortality without robust systems in place to ensure timely, accurate death reporting (7–10). In Zambia, reliance on paper-based methods leads to delays, incomplete records, and lost forms, ultimately undermining mortality surveillance (9, 11).

Timely death reporting is particularly important for detecting mortality trends and informing policies aimed at improving health outcomes. The World Health Organization (WHO) recommends the use of mobile technologies to standardize mortality reporting and improve data completeness in countries with weaker CRVS systems (12). WHO has also issued guidance on classifying digital health interventions to address specific health system challenges.

In Zambia, the Department of National Registration, Passport and Citizenship (DNRPC) under the Ministry of Home Affairs and Internal Security (MoAHIS), manages all CRVS activities. The DNRPC currently operates a manual paper-based system which supports civil registration services, including national ID issuance and birth and death registration. Data is then entered into an Integrated National Registration and Identification System (INRIS) that is deployed in select provinces. With partner support, there are plans to extend INRIS coverage to improve service accessibility and CRVS efficiency.

The technologies applied in this case study fall under System Category 4.0: Data Services, addressing Health System Challenge Category 1 (information-related challenges), such as delayed reporting, poor data quality, limited data access, and underutilization of available data (13, 14).

Despite progress, challenges persist. Death notification processes in many areas remain manual (9, 11) which delays processing and reduce the availability of real-time data for decision making. Families often must travel long distances to report a death or birth. Currently, completed Form 13 death notifications are physically transported weekly from local offices to provincial or national headquarters for data entry into INRIS. This process is slow, costly, and inefficient, delaying the production of vital statistics reports.

To address these gaps, a mobile digital platform was implemented to capture high-quality mortality data in real time, enabling immediate submission and access for analysis. This study compares the timeliness, completeness, and accuracy of death notifications processed through the new electronic system with those reported via the existing manual method. It also highlights how digitization can streamline workflows and improve coordination between the civil registration and health sectors.

## Methods

With technical support from the Bloomberg Philanthropies Data for Health (D4H) initiative and in collaboration with the DNRPC, the project team identified and implemented key actions to address inefficiencies in death reporting. The most significant steps included:

(a) Designing an electronic, mobile-based version of the official death notification form (Form 13)
(b) Training registration and health officers in the use of the tool to support data entry, form archiving, and improved coordination with local council offices to facilitate the movement of forms.

The pilot targeted provinces with high volumes of death reports and significant backlogs of unprocessed forms—namely Lusaka, Central, Copperbelt, and Southern Provinces (8, 9).

### Electronic death reporting (e-reporting)

Form 13—the official death notification document in Zambia was digitized using KoboToolbox (15) and hosted on the Ministry of Health (MoH) servers. After design and development, training sessions were conducted for DNRPC officers and district-level mortality surveillance staff to ensure smooth adoption of the system and effective data capture.

The MoH assigned mortality surveillance officers to health facilities located near mortuaries. These officers were tasked with recording all deaths, including those occurring in the facility and those brought in from the community. For community deaths, before the body is admitted to the mortuary, officers assist burial officials and the deceased’s family in completing the paper Form 13. The officer then entered the details into the digital platform, scanned supporting documents, and uploaded the cause-of-death information. Officers were required to sync their entries with the main server by the end of each day to ensure timely submission.

After completing Form 13, the family would proceed to the burial office with the form, the deceased’s documents, and any required legal paperwork. The burial officer records the death in the official register, files the documents, and issues a burial permit upon payment of the fee. This permit number is later used to obtain the official death certificate.

To address the backlog of Form 13 submissions from 2021 to 2024, district and provincial offices sent all completed forms to the DNRPC. The D4H technical team supervised and managed the allocation of these forms for data entry. At the same time, notification officers captured new deaths from 2025 while conducting verbal autopsies on community deaths (10).

The D4H team also developed an online dashboard that synced data directly with the server, enabling both field team and the DNRPC to monitor the data entry process and view results in real time.

### Dashboard Development

A central feature of the e-reporting system was the creation of a real-time death reporting dashboard. This tool served two main purposes:

1. Monitoring progress — visualizing the flow of death reports across provinces in real time.
2. Ensuring data quality — integrating built-in validation checks to automatically flag inconsistent dates, illogical age entries, or other errors at the point of data entry.

By improving accuracy and accessibility, the dashboard ensured decision-makers had timely, high-quality data for public health planning and response. It features built-in quality checks to flag inconsistent dates, implausible ages, or missing data at the point of entry. This ensures data quality and immediate accessibility for consistent public health decision-making.

### Statistical analysis

Data from the digital platform was downloaded from the server, exported to Excel, and imported into R for statistical analysis. The DNRPC also provided death data from the registration system for the years 2021-2023. The study evaluated timeliness, completeness, and accuracy of death notifications for each system. Chi-square tests were used to compare the age and sex distributions between e-reporting and traditional manual reporting records. Annual and monthly mortality trends were summarized by month, province, and age group.

Completeness rates were calculated as the proportion of total estimated deaths notified during the study period, with 95% confidence intervals. Estimated total deaths were derived from 2022 census data using crude death rates.

Timeliness was measured as the number of days from the date of death to the date of entry. Backlogged e-reporting entries were excluded to prevent bias, and only deaths reported within 30 days were analyzed. The average time to complete a digital form was also calculated. All analyses were performed in R v4.2.1.

## Findings/Results

Between July 1, 2024, and June 10, 2025, a total of 115,573 deaths were captured via the electronic notification system. Of these, 68,389 (59.2%) were male, with a median age at death of 40 years (IQR 17–62). Deaths among children under one year were the most common (18,884; 16.3%), while the 5–9 year (1,977; 1.7%) and 10–14 year (1,758; 1.5%) age groups had the fewest cases (Table 1). The age-sex distribution mirrored patterns typical of developing countries—high early childhood mortality, lower mortality during adolescence, and rising mortality between ages 20–49 (particularly among males), followed by higher female mortality in older age groups (Figure 1).

**Figure 1:**
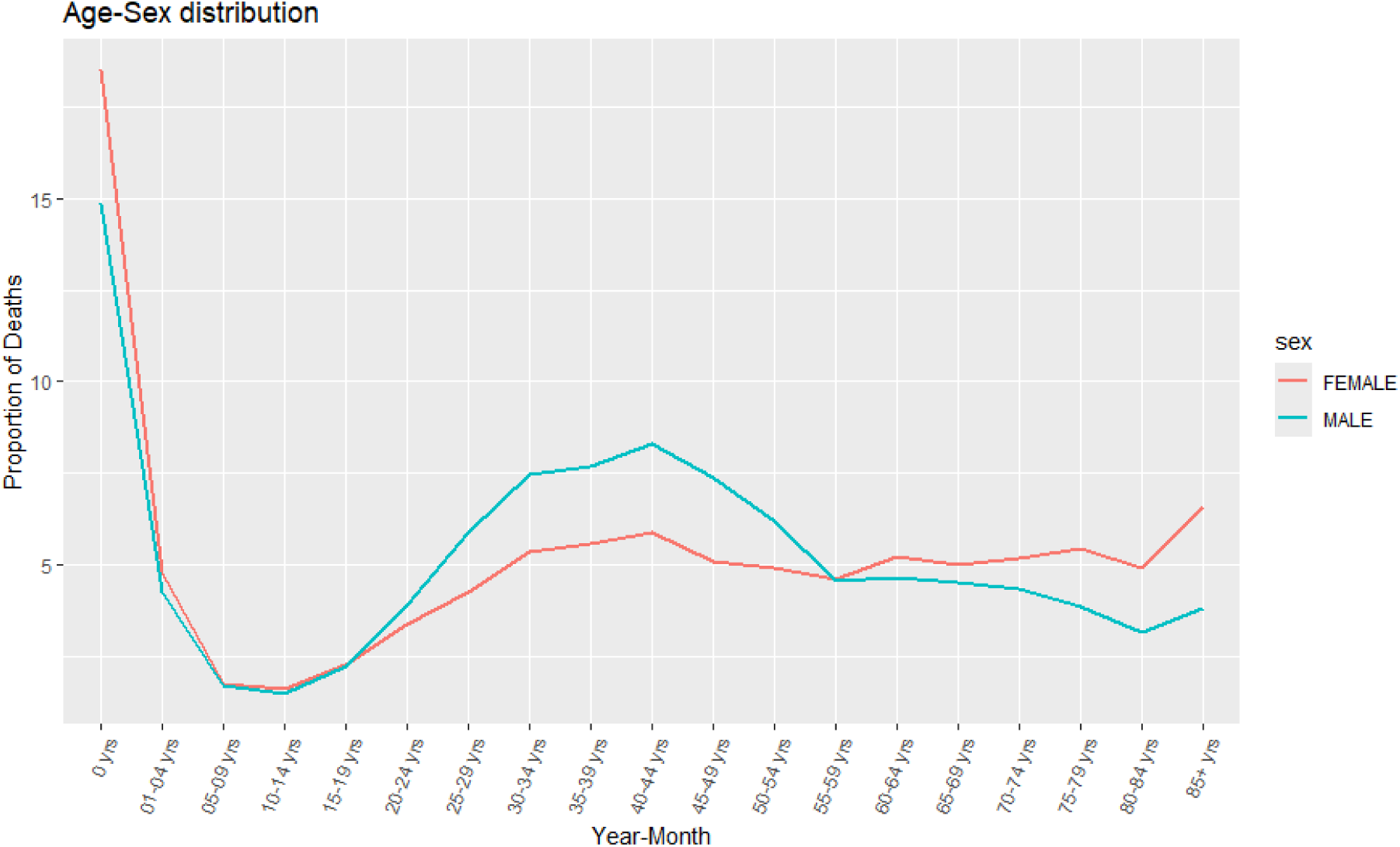
Age-Sex distribution

**Table 1:**
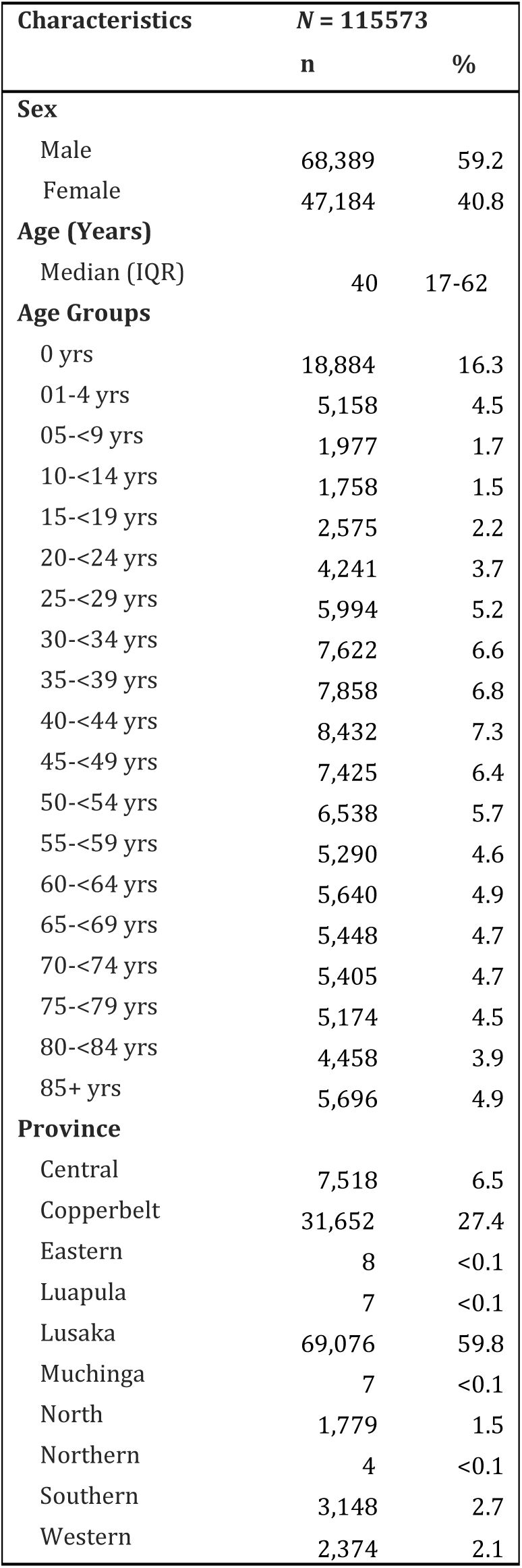
Demographic characteristics of Zambia deaths captured in e-reporting system, July 2024-June 2025.

| Characteristics | N = 115573 |  |
| --- | --- | --- |
|  | n | % |
| **Sex** |  |  |
| Male | 68,389 | 59.2 |
| Female | 47,184 | 40.8 |
| **Age (Years)** |  |  |
| Median (IQR) | 40 | 17-62 |
| **Age Groups** |  |  |
| 0 yrs | 18,884 | 16.3 |
| 01-4 yrs | 5,158 | 4.5 |
| 05-<9 yrs | 1,977 | 1.7 |
| 10-<14 yrs | 1,758 | 1.5 |
| 15-<19 yrs | 2,575 | 2.2 |
| 20-<24 yrs | 4,241 | 3.7 |
| 25-<29 yrs | 5,994 | 5.2 |
| 30-<34 yrs | 7,622 | 6.6 |
| 35-<39 yrs | 7,858 | 6.8 |
| 40-<44 yrs | 8,432 | 7.3 |
| 45-<49 yrs | 7,425 | 6.4 |
| 50-<54 yrs | 6,538 | 5.7 |
| 55-<59 yrs | 5,290 | 4.6 |
| 60-<64 yrs | 5,640 | 4.9 |
| 65-<69 yrs | 5,448 | 4.7 |
| 70-<74 yrs | 5,405 | 4.7 |
| 75-<79 yrs | 5,174 | 4.5 |
| 80-<84 yrs | 4,458 | 3.9 |
| 85+ yrs | 5,696 | 4.9 |
| **Province** |  |  |
| Central | 7,518 | 6.5 |
| Copperbelt | 31,652 | 27.4 |
| Eastern | 8 | <0.1 |
| Luapula | 7 | <0.1 |
| Lusaka | 69,076 | 59.8 |
| Muchinga | 7 | <0.1 |
| North | 1,779 | 1.5 |
| Northern | 4 | <0.1 |
| Southern | 3,148 | 2.7 |
| Western | 2,374 | 2.1 |

Spatially, Lusaka Province accounted for the majority of reported deaths (59.8%). On average, completing a digital form took 4.6 minutes (median 4.0, IQR 2.9–5.7 minutes).

### Comparison between deaths captured through manual paper-based system and deaths captured through electronic reporting system

#### Distribution of deaths

A significantly higher number of deaths were recorded by the electronic reporting system compared with those captured by the manual system submitted through INRIS (χ2(1) =4.01, *p=<0.045*) (Figure 2).

**Figure 2:**
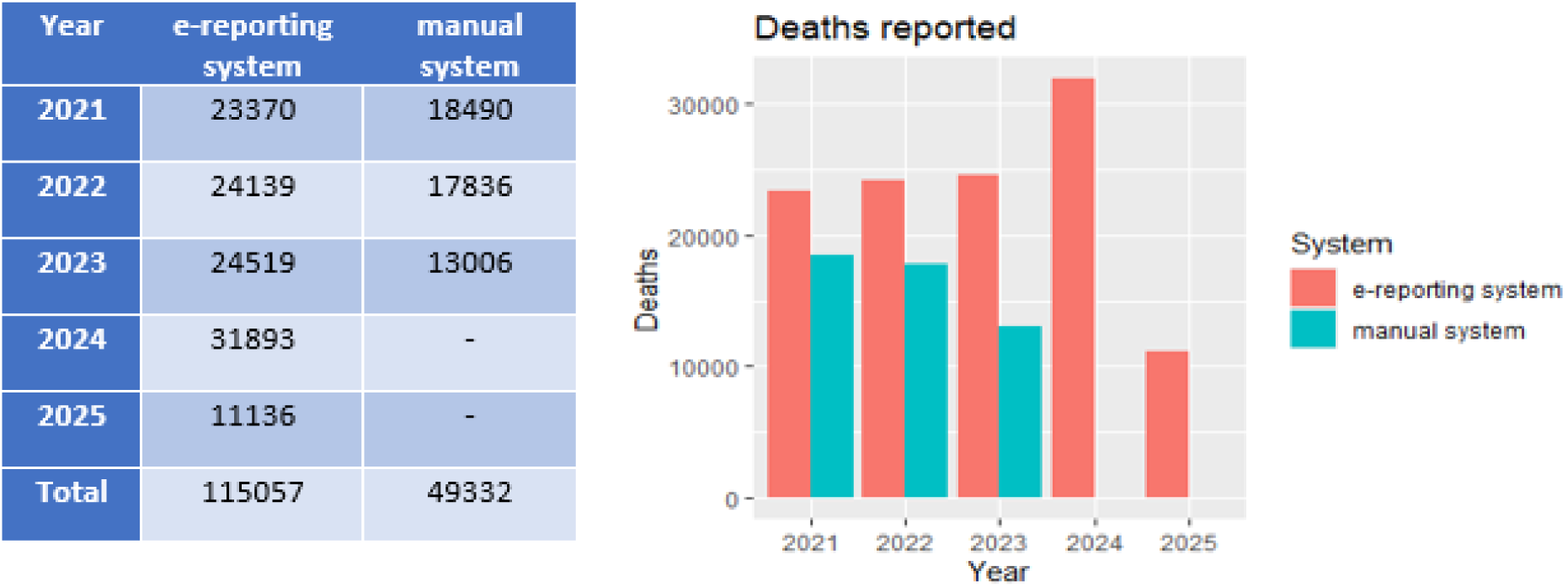
Number of deaths captured through the manual system and e-reporting system.

Both systems showed high mortality in early childhood, a decline during childhood and adolescence, followed by an increase between ages 20-49 and a subsequent decline to more stable levels in older age groups (Figure 3).

**Figure 3:**
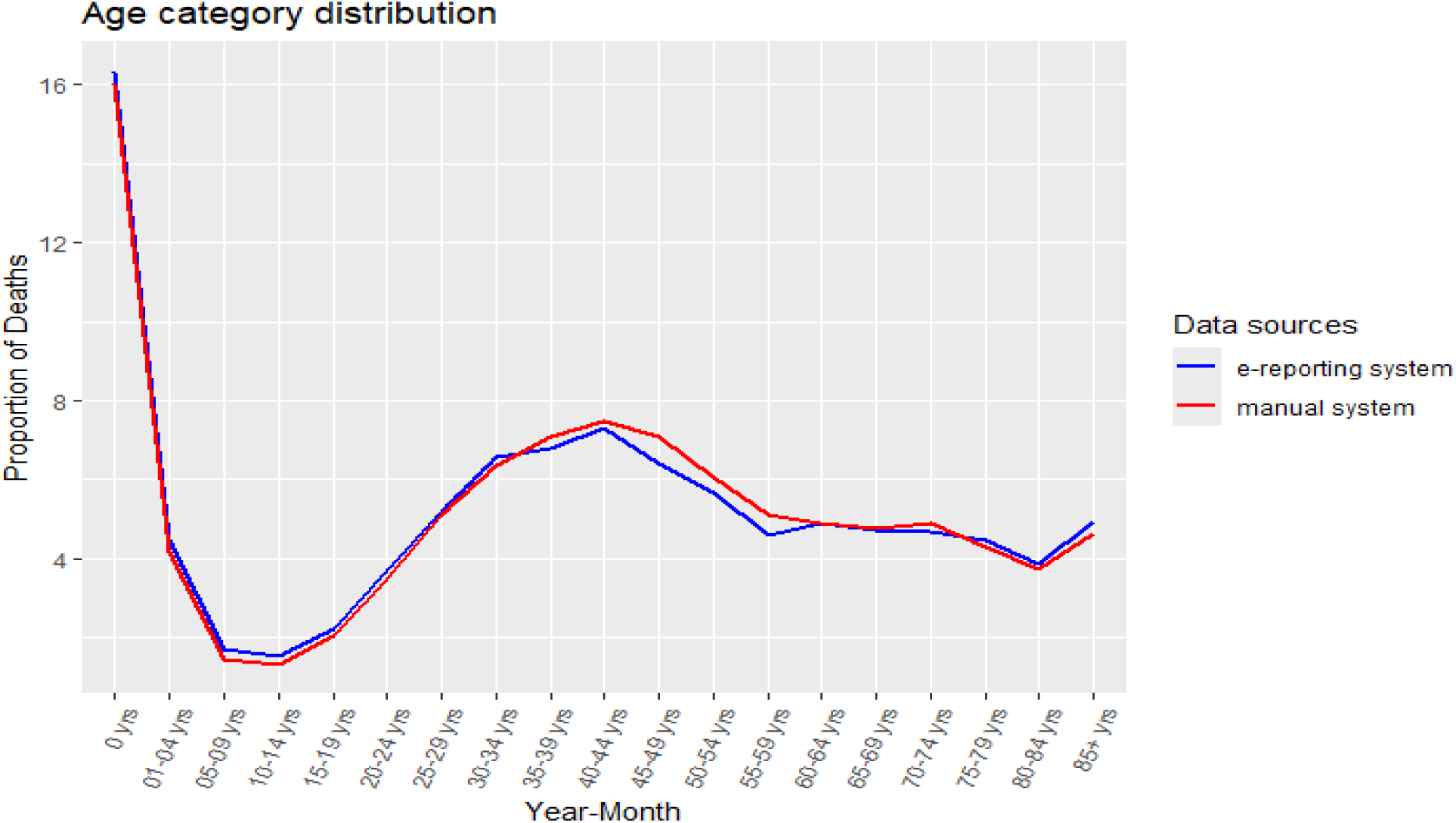
Age category distribution from the manual system and e-reporting system.

#### Data quality of Age values between both systems

When assessing age-related data quality, we found that the manual system contained substantial outliers, including ages over 250 years, revealing significant data entry errors. The e-reporting system, however, demonstrated a marked improvement in data quality, largely because it was designed with enhanced quality checks integrated directly into the data entry process. Additionally, the e-reporting system proved more comprehensive, capturing a wider range of data (IQR=45) with a lower probability of missing information in specific age groups compared to the manual system through INRIS (IQR=42) (Figure 4). Ultimately, the improved quality and constant availability of data from the e-reporting system leads to greater efficiency.

**Figure 4:**
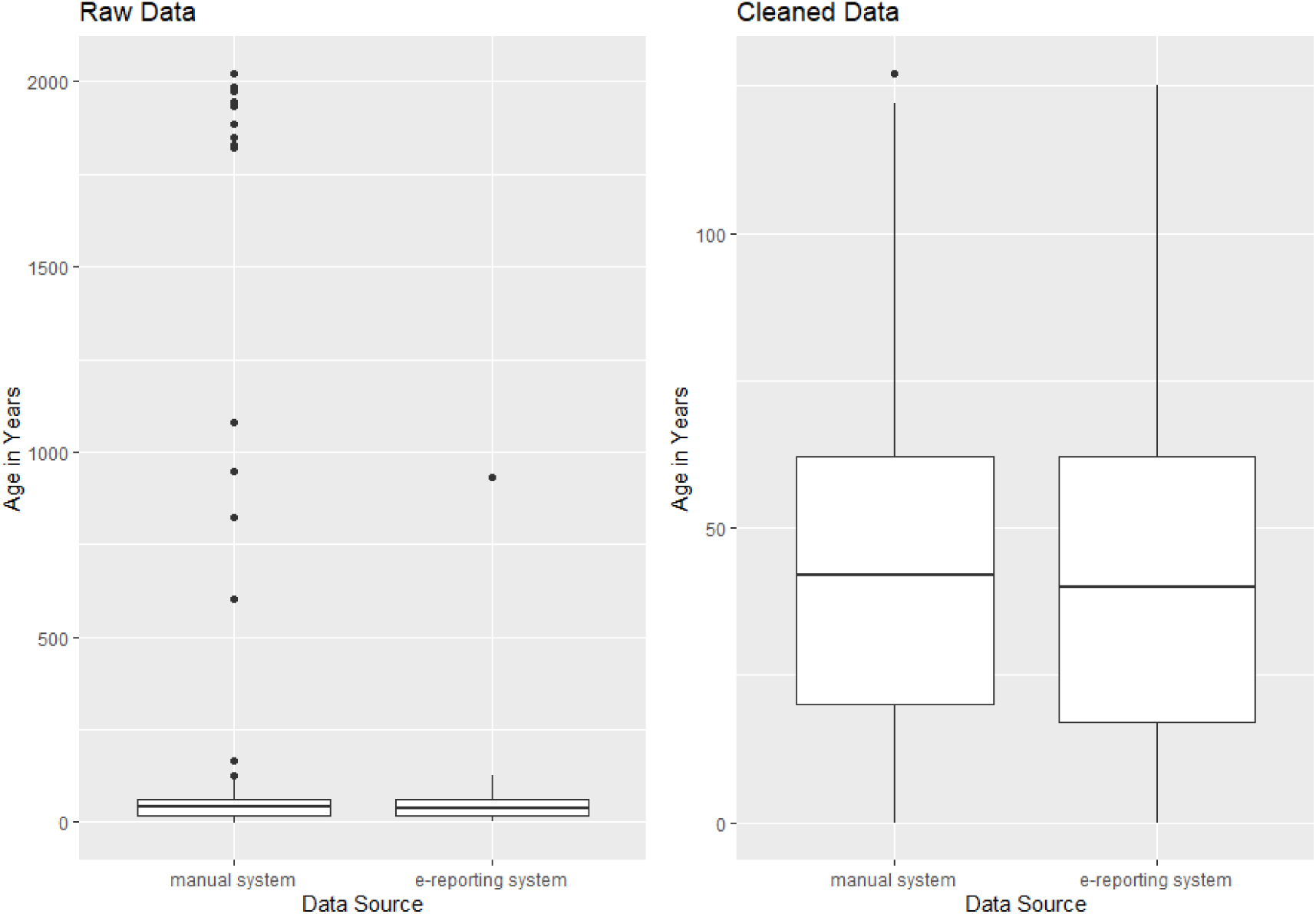
Data Quality in Age (Original data) and Data Quality in Age (Cleaned data)

#### Completeness and Timeliness

The introduction of the e-reporting system was primarily driven by a need to address long-standing issues of completeness and timeliness in Zambia’s death notification and registration process. Analysis comparing the new system with the manual system demonstrates a significant improvement in both areas.

Regarding completeness, the e-reporting system captured a higher proportion of deaths, achieving an estimated rate of 17.4% (95% CI: 17.3%-17.5%). This marks a notable improvement over the manual system, which recorded a lower completeness rate of 11.9% (95% CI: 11.8%-12.0%).

For timeliness, the e-reporting system also proved to be more efficient. The average reporting time from the date of death was approximately 3 days, with a variance of 24 days (Figure 5). This represents a significant reduction compared to the manual system, which had an average reporting time of 6 days and a much larger variance of 62 days. Statistical analysis confirmed that this difference is highly significant (p < 0.001), highlighting the e-reporting system’s effectiveness in accelerating the death notification process.

**Figure 5:**
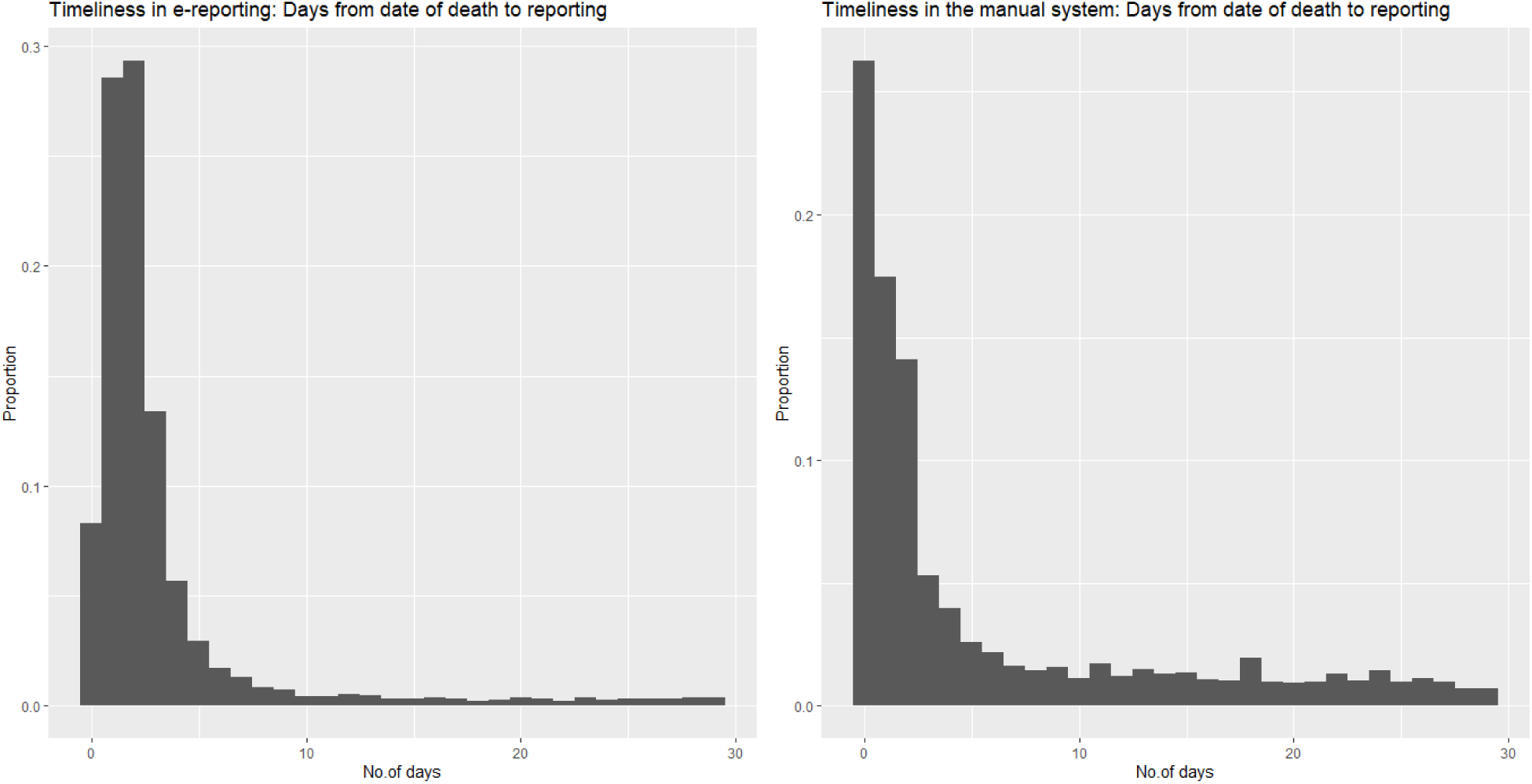
Reporting Timeliness comparison between e-reporting system and the manual system.

## Discussion

The civil registration system in Zambia, like those in many developing nations, faces significant challenges with robust and systematic death reporting. This study highlights the importance of integrating electronic data collection and analysis platforms into CRVS systems to dramatically improve completeness and timeliness. The e-reporting system provides a platform that delivers more reliable, accurate, and timely death reporting. We observed a notable increase in reported deaths using this system—a finding consistent with similar initiatives in other developing countries. For example, electronic notification has proven effective in nations such as Papua New Guinea and Tanzania, where it increased death notifications and reduced reporting delays (5).

A key factor in the system’s success is its ability to facilitate timely death reporting and data collection. The mobile digital platform ensures data is captured in real-time, maintains high data quality, and submits data to the server daily, thus reducing delays in the notification process. We found that the electronic platform significantly outperformed the manual process in speed. The developed online dashboard, which periodically syncs data from the server, further supports this by providing crucial monitoring and data summaries for public health surveillance. The World Health Organization (WHO) emphasizes the use of dashboards and digital tools as a key component of modern CRVS strengthening, and our findings align with this recommendation (12).

## Limitations

This study has several limitations. First, data collection was centralized in select provinces where D4H staff are located. Consequently, the findings may not be fully representative of the entire country. This intervention was designed to support the limited workforce at the DNRPC, which was struggling with existing workloads. The results underscore the importance and significance of implementing such platforms in underdeveloped CRVS systems. Second, the e-reporting system has not yet been integrated into the main CRVS system, although discussions are underway. Full integration would likely enhance the death registration process and further improve the CRVS system. Third, the study relied on passive reporting from families visiting the burial office, rather than real-time community-level death reporting. This means some deaths in the community may still be missed, underscoring the need for a more proactive reporting model to improve coverage.

## Conclusion

The implementation of the e-reporting system has successfully enhanced the tracking of death notifications. Periodic assessment of these notifications is critical in ensuring that all deaths are reported and registered without duplication, information is accurately recorded, and timeliness is maintained. The introduction of this electronic system has proven to be an effective strategy for promptly capturing high-quality death reports. This not only reduces the chances of missing deaths but also improves overall death reporting coverage, ultimately enhancing our ability to track disease trends, assess the impact of health programs, and respond effectively to public health emergencies. This initiative represents a significant step towards strengthening Zambia’s CRVS system.

## Acknowledgement

We appreciate all the D4H registrars and MOH mortality surveillance officers who dedicated their time to support data entry and the DNRPC staff for providing Form 13 for the data entry exercise. We also thank the Ministry of Home Affairs through the Department of National Registration, Passports and Citizenship (DNRPC), for allowing us to use this data and the CDC Foundation and Bloomberg Philanthropies D4H initiative for project support.

## Funding

Financial support was provided by the Bloomberg Philanthropies Data for Health Initiative through the CDC Foundation with a grant from Bloomberg Philanthropies.

## Data availability

The datasets generated and/or analysed during the current study are not publicly available due to the sensitivity of the data; however, they may be made available upon reasonable request from the Ministry of Home Affairs through official channels of the Government of Zambia and the Department of National Registration, Passports and Citizenship (DNRPC).

## Potential Conflict of Interest

The contents of this manuscript are those of the authors and do not necessarily represent the official position of the CDC Foundation and Bloomberg Philanthropies.

The authors declare there is no conflict of interest in relation to the publication of this work.

## Notes

### Competing Interest Statement

The authors have declared no competing interest.

### Author Declarations

This is a Civil Registration and Vital Statistics (CRVS) program which is a government-mandated national system implemented on a routine basis. Data was obtained with formal authorization from the Department of National Registration, Passport, and Citizenship (DNRPC) as part of routine CRVS operations. As the work focused on strengthening CRVS death reporting and notification processes, Institutional Review Board (IRB) approval was not required. To protect privacy and confidentiality, all data extracted from the CRVS system were fully de-identified prior to analysis, with no personally identifiable information retained, in accordance with ethical standards for the secondary use of public health data.

